# Cerebrospinal fluid hemoglobin drives subarachnoid hemorrhage-related secondary brain injury

**DOI:** 10.1101/2021.02.13.21251469

**Authors:** Kevin Akeret, Raphael M. Buzzi, Christian A. Schaer, Bart R. Thomson, Florence Vallelian, Sophie Wang, Jan Willms, Martina Sebök, Ulrike Held, Jeremy W. Deuel, Rok Humar, Luca Regli, Emanuela Keller, Michael Hugelshofer, Dominik J. Schaer

## Abstract

Secondary brain injury after aneurysmal subarachnoid hemorrhage (SAH-SBI) is a significant contributor to poor outcomes in patients after rupture of an intracranial aneurysm. The lack of diagnostic biomarkers and novel drug targets represent an unmet need. Prior experimental evidence has suggested cell-free hemoglobin in the cerebrospinal fluid (CSF-Hb) as a pathophysiological driver of SAH-SBI. The aim of this study was to investigate the clinical and pathophysiological association between CSF-Hb and SAH-SBI. We prospectively enrolled 47 consecutive patients and collected daily CSF samples within 14 days after aneurysm rupture. There was very strong evidence for a positive association between CSF-Hb and SAH-SBI. The diagnostic accuracy of CSF-Hb for SAH-SBI markedly exceeded that of established methods (area under the curve: 0.89 [0.85-0.92]). Temporal LC-MS/MS CSF proteomics demonstrated that erythrolysis accompanied by an adaptive macrophage response are the two dominant biological processes occurring in the CSF space after aneurysm rupture. To further investigate the pathophysiology between CSF-Hb and SAH-SBI, we explored the vasoconstrictive and lipid peroxidation activities of Hb ex-vivo. These experiments revealed critical inflection points overlapping CSF-Hb concentration thresholds in patients with SAH-SBI. Selective Hb depletion and in-solution neutralization by the Hb-scavenger haptoglobin or the heme-scavenger hemopexin efficiently attenuated the vasoconstrictive and lipid peroxidation activities of CSF-Hb in patient CSF. Collectively, the clinical association between high CSF-Hb levels and SAH-SBI, the underlying pathophysiological rationale, and the favorable effects of haptoglobin and hemopexin in ex-vivo experiments position CSF-Hb as a highly attractive biomarker and potential drug target.

## INTRODUCTION

Aneurysmal subarachnoid hemorrhage (aSAH) accounts for only 5-10% of all strokes *(1)*, but the associated morbidity and socioeconomic burden exceed those of ischemic strokes due to the younger age of affected patients *(2)*. In addition to early brain injury within the first 72 hours *(3)*, patient outcomes are determined by delayed secondary brain injury (SAH-SBI), which often occurs between days 3 and 14 after aneurysm rupture *(4)*. Two-thirds of patients develop angiographic vasospasm (aVSP) in the large cerebral arteries *(5)*. Delayed cerebral ischemia (DCI) with radiologic demarcation of ischemic brain areas and clinically evident delayed ischemic neurologic deficits (DINDs) are found in one-third of patients with aSAH *(6)*. The pathophysiology of SAH-SBI is multifactorial, involving macro- and microvascular dysfunction, neuroinflammation, neuronal apoptosis, and pathological electrical brain activity *(7–9)*. The delay between aneurysm rupture and the onset of SAH-SBI provides a window of opportunity for preventive and therapeutic interventions. However, to date, the only preventive intervention for SAH-SBI that has been shown to moderately improve neurological outcomes is oral nimodipine *(10, 11)*. In already symptomatic patients, the therapeutic options are limited to noncausal rescue therapies to improve cerebral perfusion, such as the adaptation of blood pressure targets or angioplasty *(10, 11)*. Therefore, methods to identify patients at high risk for SAH-SBI as well as novel therapeutic targets represent an unmet need.

Based on preclinical studies *(12–15)*, we propose the following pathophysiological sequence to underlie the increased risk for SAH-SBI: red blood cells (RBCs) decompose in the subarachnoid hematoma, cell-free Hb is generated in the CSF (CSF-Hb), Hb tetramers dissociate into dimers, and finally small CSF-Hb dimers delocalize into vulnerable anatomical sites of the cerebral arteries and brain. There, nitric oxide (NO) scavenging promotes vasoconstriction *(16, 17)* and potentially oxidative neuronal damage. In experimental models, these adverse effects of CSF-Hb were mitigated by intraventricular administration of the Hb scavenger protein haptoglobin *(16)*. Collectively, this prior evidence suggests CSF-Hb as a biomarker that may allow the risk of SAH-SBI to be estimated by directly monitoring a potentially targetable pathophysiological process.

The main objective of this study was to investigate the clinical association of CSF-Hb with SAH-SBI and the diagnostic accuracy of daily CSF-Hb measurements. Furthermore, we performed an in-depth CSF proteome study and ex vivo functional assays to rationalize the role of CSF-Hb as a pathophysiological driver of and potential therapeutic target for SAH-SBI.

## RESULTS

### Study population

Out of 52 consecutively screened patients fulfilling the eligibility criteria, 47 were included, since for five patients informed consent was refused. Table 1 summarizes the baseline features of the patient cohort.

**Table 1.**
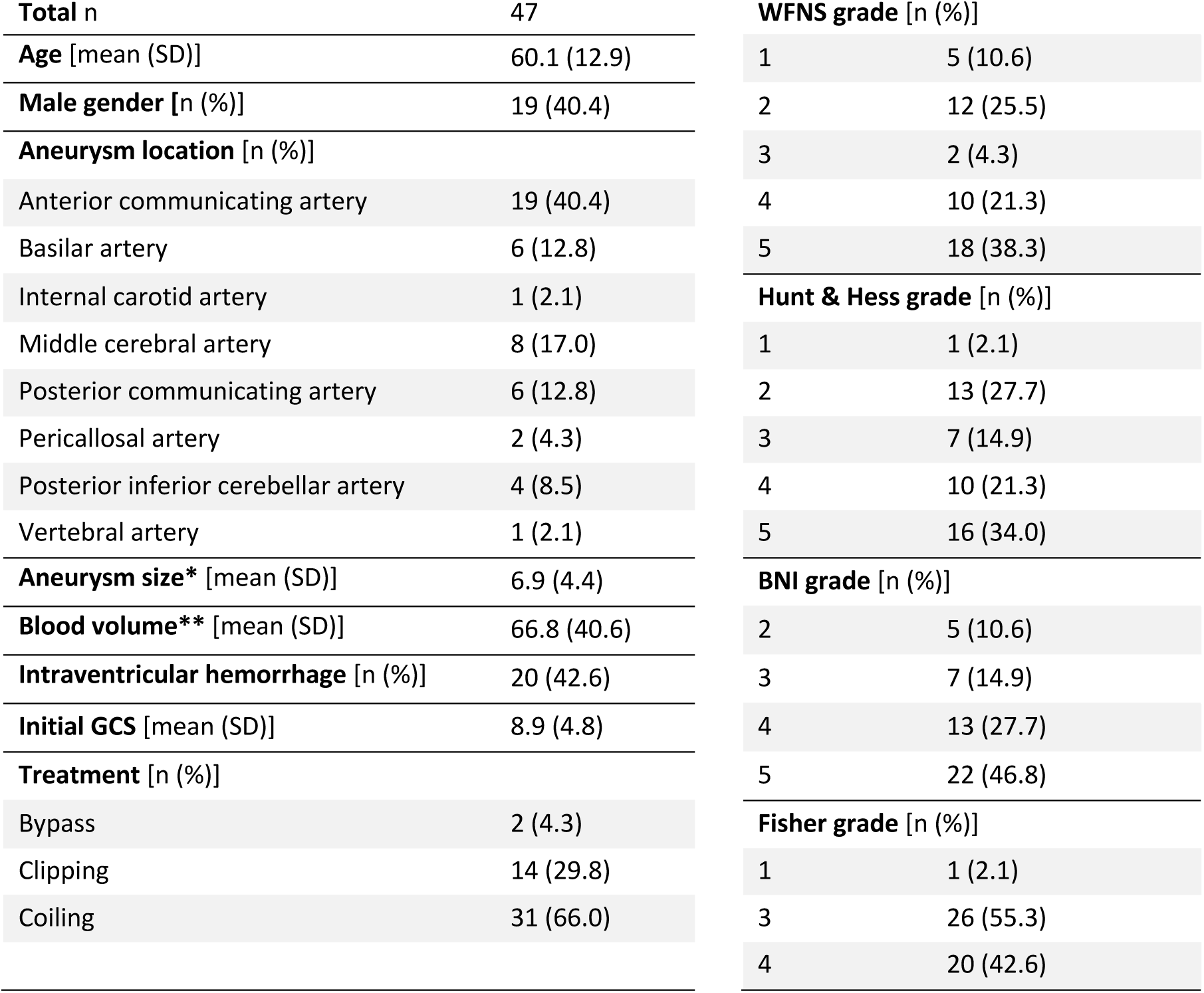
Baseline cohort characteristics. Demographic, clinical and radiological cohort features at baseline are shown. *BNI = Barrow Neurological Institute; GCS = Glasgow Coma Scale; SD = standard deviation; WFNS = World Federation of Neurosurgical Societies. * maximal diameter in mm; ** volume in cm*^*3*^

### Spectrophotometry demonstrates delayed accumulation of CSF-Hb and heme metabolites in patient CSF

We used spectrophotometry with spectral deconvolution to quantify CSF-Hb, its downstream metabolites biliverdin and bilirubin, and the primary Hb oxidation product metHb (Fig. 1A). The individual temporal profiles and peak concentrations of CSF-Hb (0.6 µM to 242.2 µM) were highly variable (Supplementary Fig. 2). In most patients, CSF-Hb remained very low over the first 2 to 3 days after aSAH, followed by a strong increase, a plateau between days 9 and 12, and a decrease thereafter. Bilirubin was already elevated on day 1 and increased at the greatest rate within the first 3 to 5 days before it reached a plateau. Levels of the intermediate metabolite biliverdin markedly increased from day 4 and peaked on day 12. MetHb levels showed a delayed increase with a peak on day 11 after aSAH. Hence, high levels of CSF-Hb develop with a delay after the acute bleeding and coincide with the high-risk phase for SAH-SBI.

**Fig. 1.**
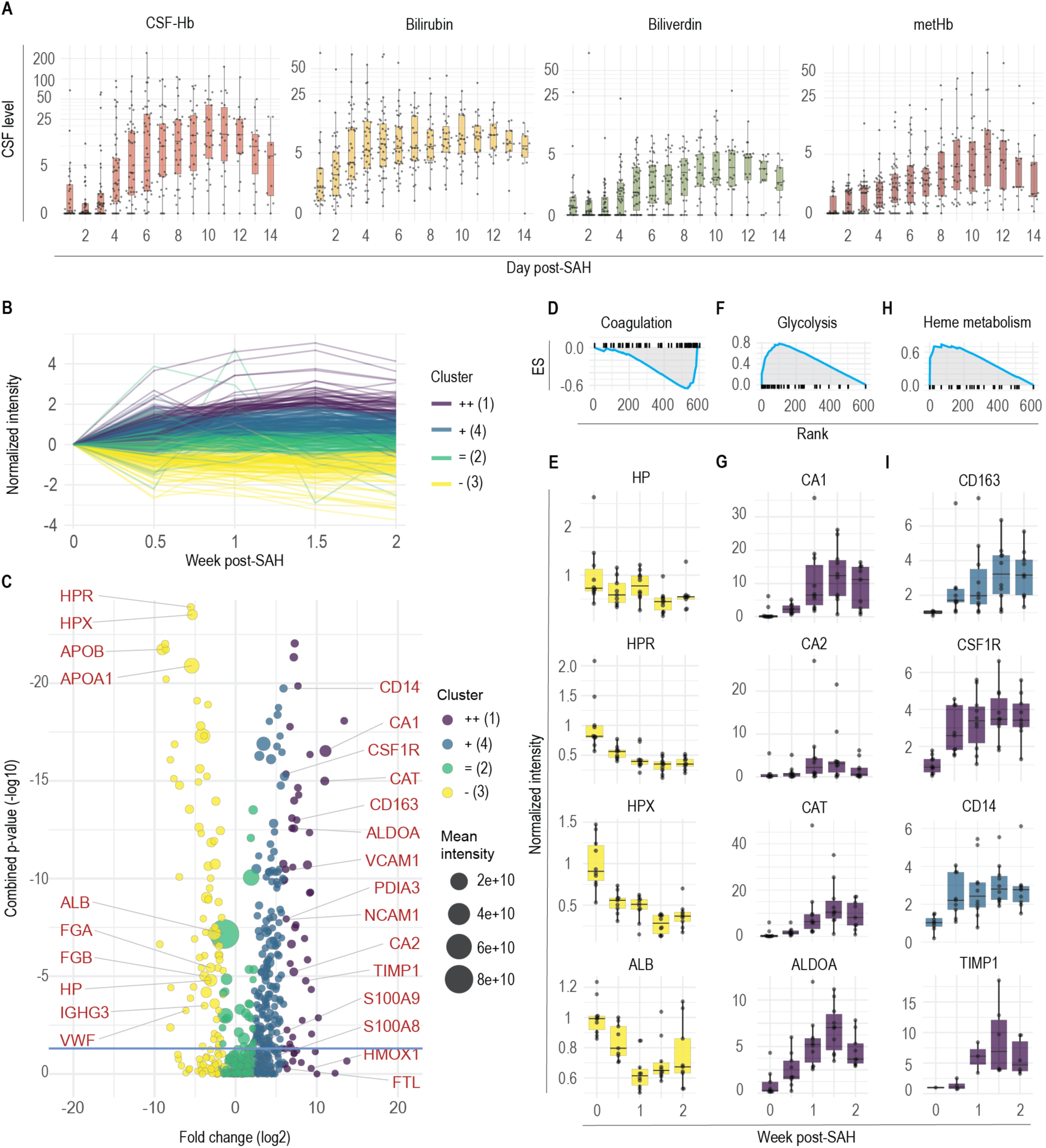
Changes in the cerebrospinal fluid proteome after aneurysmal subarachnoid hemorrhage. **A**. Temporal cerebrospinal fluid (CSF) oxyhemoglobin (CSF-CSF-Hb), bilirubin, biliverdin and methemoglobin (metHb) profiles. **B**. K-means clustering of proteins in the CSF after aneurysmal subarachnoid hemorrhage (aSAH) identified with LC-MS/MS. **C**. Volcano plot showing the overall fold change and combined p-value for the CSF proteome. The color indicates the respective cluster, the size of each dot represents the raw mean intensity of the protein (not normalized). **D**. Enrichment plot showing the top negatively enriched hallmark gene sets for coagulation identified by gene set enrichment analysis (GSEA) of the CSF proteome after aSAH. **E**. Temporal course of the normalized protein intensities for haptoglobin (HP), haptoglobin-related protein (HPR), hemopexin (HPX) and albumin (ALB) in the CSF after aSAH. **F**. Enrichment plot of the top positively enriched hallmark gene set for glycolysis identified by GSEA of the CSF proteome after aSAH. **H**. Enrichment plot of the hallmark gene set for heme metabolism identified by GSEA of the CSF proteome after aSAH. **G**. Temporal course of the normalized protein intensities for carbonic anhydrase 1 (CA1), carbonic anhydrase 2 (CA2), catalase (CAT) and aldolase A (ALDOA) in the CSF. **I**. Temporal course of the normalized protein intensities for CD163, colony stimulating factor 1 receptor (CSF1R), CD14 and metalloproteinase inhibitor 1 (TIMP) in the CSF. *The box in the boxplots bounds the interquartile range (IQR) divided by the median, while the whiskers extend to the highest and lowest value within the 1*.*5 x IQR, respectively*.

**Fig. 2.**
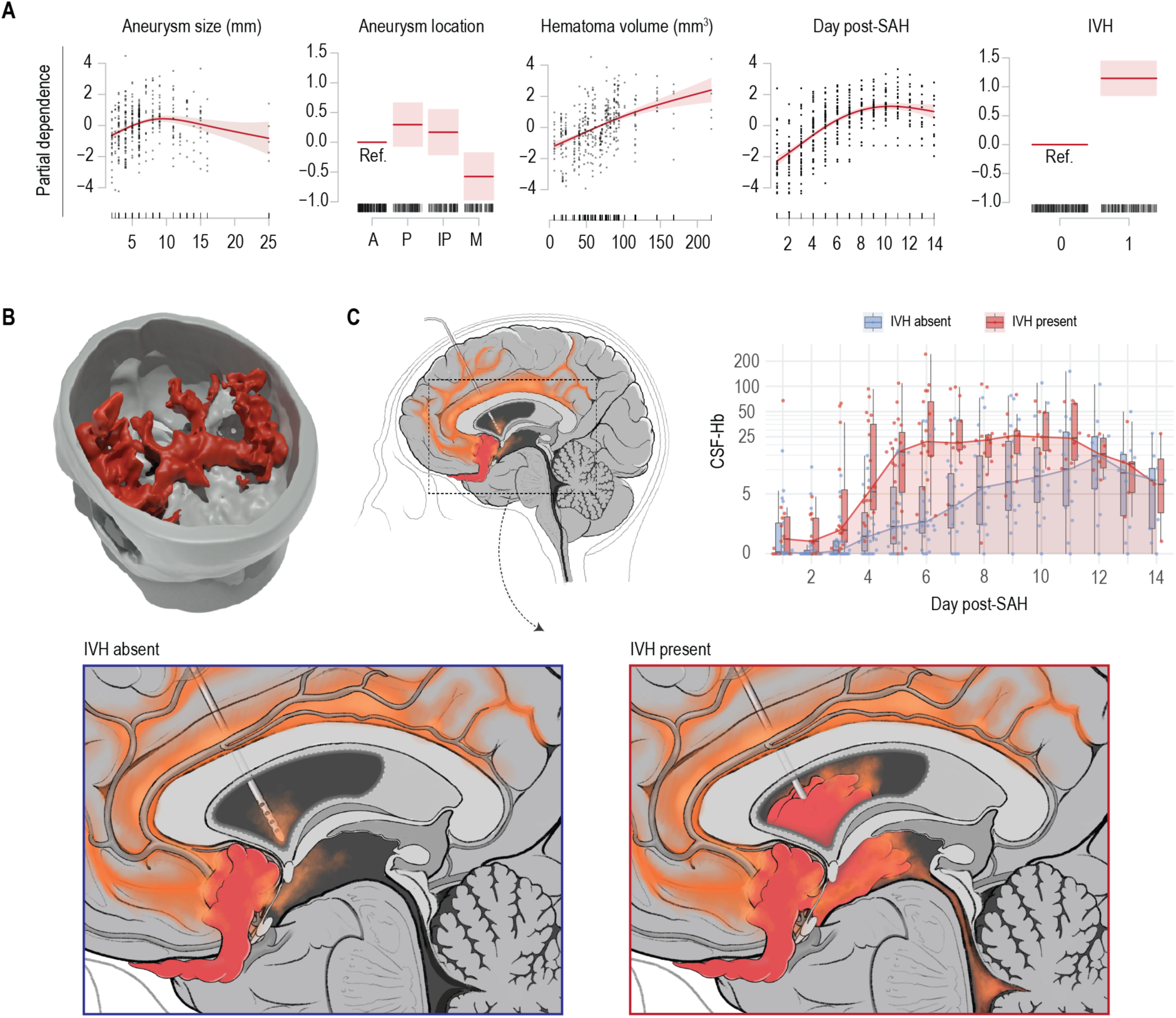
Determinants of cerebrospinal fluid hemoglobin. **A**. Partial dependence of cerebrospinal fluid hemoglobin (CSF-Hb) levels on aneurysm size, aneurysm location, hematoma volume, the presence of intraventricular hemorrhage (IVH), and number of days post-SAH according to a generalized additive model (GAM). **B**. Example of 3D-rendered subarachnoid hematoma. **C**. *Left:* Schematic illustration of a subarachnoid hemorrhage originating from a ruptured anterior communicating artery aneurysm with IVH absent (blue inset) or present (red inset). *Right:* The temporal course of CSF-Hb after aSAH stratified by the presence of IVH. *The box in the boxplots bounds the interquartile range (IQR) divided by the median, while the whiskers extend to the highest and lowest value within the 1*.*5 x IQR, respectively*.

### LC-MS/MS analysis of CSF proteins delineates erythrolysis and a dynamic macrophage response in the subarachnoid space

We performed a quantitative LC-MS/MS analysis of 85 CSF samples from 18 patients collected at 5 time points after aSAH. Figure 1B illustrates the temporal course of the levels of 757 CSF proteins that were assigned to four clusters by a k-means algorithm. Figure 1C shows a volcano plot of the normalized signal intensities summed across all samples per patient, providing an overall view of protein accumulation or depletion within the two-week period after aSAH. Cluster 2 (green) comprised proteins whose intensity remained mostly unchanged over time. Cluster 3 (yellow) represents proteins with decreasing intensity over time. These were mainly plasma proteins that likely entered the subarachnoid space with the bleeding and were subsequently cleared from the CSF. GSEA of the ranked proteins assigned the top negatively enriched hallmark gene set to coagulation (ES = −0.66, FDR = 0.007, Fig. 1D). Figure 1E displays the levels of four selected plasma proteins (HP, HPR, HPX and ALB) over time. Cluster 1 (violet) contained proteins that showed pronounced accumulation over time, and cluster 4 (blue) contained proteins whose levels showed a moderate increase. GSEA identified glycolysis (ES = 0.78, FDR = 0.021, Fig. 1F) and heme metabolism (ES = 0.74, FDR = 0.096, Fig. 1H) as the top enriched hallmark gene sets, indicating the lysis of RBCs with subsequent release of cytoplasmic proteins and the breakdown of Hb by macrophages, respectively. Figure 1G displays levels of the RBC proteins carbonic anhydrase 1 (CA1) and 2 (CA2), catalase (CAT), and aldolase A (ALDOA) over time. Figure 1I shows the macrophage proteins CD163, CSF1R, CD14, and TIMP-1. The accumulation of these proteins suggests a pronounced macrophage influx into the subarachnoid space within the first days after hemorrhage. Collectively, the proteome dynamics in patient CSF after aSAH reinforced the notion that erythrolysis and an adaptive macrophage response are the two dominant processes in the CSF space after aSAH.

### Hematoma volume, day after aSAH and presence of IVH as relevant determinants of CSF-Hb

To estimate determinants of CSF-Hb concentration, we applied a GAM with non-linear spline-fit (4 knots) for time (day after aSAH) and a random-effect for the individual patients. The partial dependence plots are shown in Figure 2A. Larger aneurysm size was not associated with higher CSF-Hb. Among aneurysms at different locations, middle cerebral artery aneurysms showed strong evidence for a negative association with CSF-Hb (partial dependence = −0.57, SD = 0.20, p = 0.0044). There was very strong evidence for a positive association between hematoma volume and CSF-Hb (p < 0.001). A rendered example of the subarachnoid blood clot is given in Figure 2B. Volumetric analysis was performed in only 46 of the 47 patients, as one patient had an initial MRI (and not a CT scan) and was therefore excluded from this analysis. There was very strong evidence (p < 0.001) for an association between the time point of sampling (day) and CSF-Hb, whereas the partial dependence was negative in the first 3-5 days after aSAH, and positive in subsequent days. This association most likely reflects the delayed onset of erythrolysis and CSF-Hb liberation in the subarachnoid space after aSAH. Also, there was very strong evidence for a positive association between the presence of IVH and CSF-Hb (partial dependence = 1.1, SD = 0.15, p < 0.0001). With IVH, CSF-Hb levels increased early and then reached a plateau, whereas in the absence of IVH, this increase was more delayed and gradual, but a plateau was reached at comparable peak concentrations around day 12 (Fig. 2C). This most likely represents a CSF compartment effect, since the EVD used to sample the CSF is in direct proximity to the blood clot when IVH is present, while the EVD lies distant to the blood clot, if IVH is absent (illustrated in Fig. 2C). In summary, these findings indicate that hematoma volume, day after aSAH and the presence of IVH are relevant determinants of CSF-Hb levels.

### Increased CSF-Hb levels are associated with SAH-SBI

The cohort outcome parameters representative of SAH-SBI, rescue therapies, complications, mortality, and functional status at the 3-month follow-up are summarized in Supplementary Table 2. The CSF-Hb levels stratified by aVSP, DCI, DIND and SAH-SBI are shown in Figure 3A and Supplementary Figure 3. As expected, given their strict clinical definitions, the status of DIND, DCI and aVSP is missing on many time points (denoted as not available (NA)). Supplemental Fig. 3 shows the CSF-Hb values of the NAs for DINDs, DCI and aVSPs compared to the CSF-Hb. Within the NAs, the CSF-Hb values were found distributed along the entire measurement spectrum without an unidirectional trend, as opposed to those with or without DIND, DCI or aVSP. A GAM with spline fit (5 knots) for time (day after aSAH) and a random-effect for the individual patients provided very strong evidence (p < 0.001) for a positive association between CSF-Hb and SAH-SBI. The partial dependence of SAH-SBI on CSF-Hb and the day after aSAH are given in Figure 3B.

**Fig. 3.**
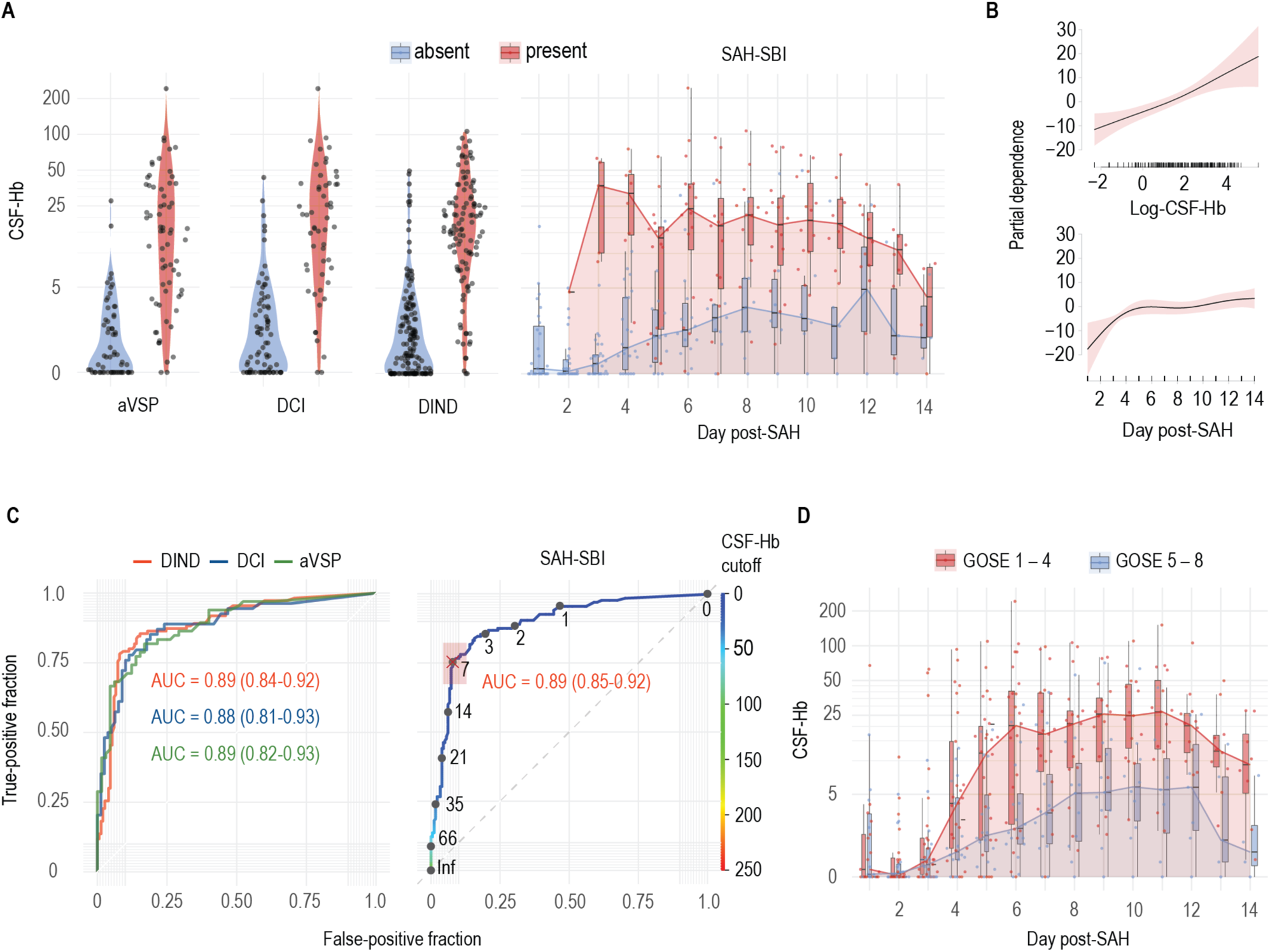
Association between cerebrospinal fluid hemoglobin and secondary brain injury after aneurysmal subarachnoid hemorrhage. **A**. Cerebrospinal fluid hemoglobin (CSF-Hb) in patients after aneurysmal subarachnoid hemorrhage (aSAH) stratified by angiographic vasospasm (aVSP), delayed cerebral ischemia (DCI), delayed ischemic neurological deficit (DIND) and the composite outcome SAH-related secondary brain injury (SAH-SBI) per day after aSAH (day post-SAH). **B**. Partial dependence of SAH-SBI on CSF-Hb (log-scale) and the number of days post-SAH according to a generalized additive model (GAM). **C**. Receiver operating characteristic (ROC) curves and area under the curves (AUC) of CSF-Hb for DIND, DCI and aVSP (left). ROC curve of CSF-Hb for SAH-SBI, with the corresponding CSF-Hb measurements. **D**. The temporal course of CSF-Hb stratified by GOSE score at the 3-month follow-up. *The box in the boxplots bounds the interquartile range (IQR) divided by the median, while the whiskers extend to the highest and lowest value within the 1*.*5 x IQR, respectively*.

To estimate the diagnostic accuracy of daily CSF-Hb measurements to monitor for SAH-SBI, we calculated the ROC curves and AUCs for SAH-SBI and the individual outcomes aVSP, DCI and DIND (Fig. 3C, Supplementary Fig. 4A-C). The high diagnostic accuracy of CSF-Hb remained unaltered in a recalculated model with data limited to the high-risk period (days 4-14), avoiding potential bias introduced by the generally low Hb levels in the first 3 days after aSAH (Supplementary Fig. 4D). Bilirubin, biliverdin and metHb had a lower diagnostic accuracy than CSF-Hb (Supplementary Fig. 4E-G). Computation of the optimal Youden index yielded a CSF-Hb value of 7.1 µM for SAH-SBI (Fig. 3C). For the individual outcomes of aVSPs, DCI and DINDs, the optimal Youden indices were 3.4, 5.5, and 7.1 µM, respectively (Supplementary Fig. 4A-C). TCD measurements demonstrated a high specificity for SAH-SBI (0.97 [0.88 to 1.00]), DIND (0.96 [0.87 to 1.00]), DCI (0.93 [0.76 to 0.99]) and aVSP (0.96 [0.78 to 1.00]). However, the sensitivity of daily TCD measurements was minimal (SAH-SBI: 0.28 [0.17 to 0.42], DIND: 0.32 [0.18 to 0.48], DCI: 0.15 [0.03 to 0.38], aVSP: 0.17 [0.05 to 0.37]).

**Fig. 4.**
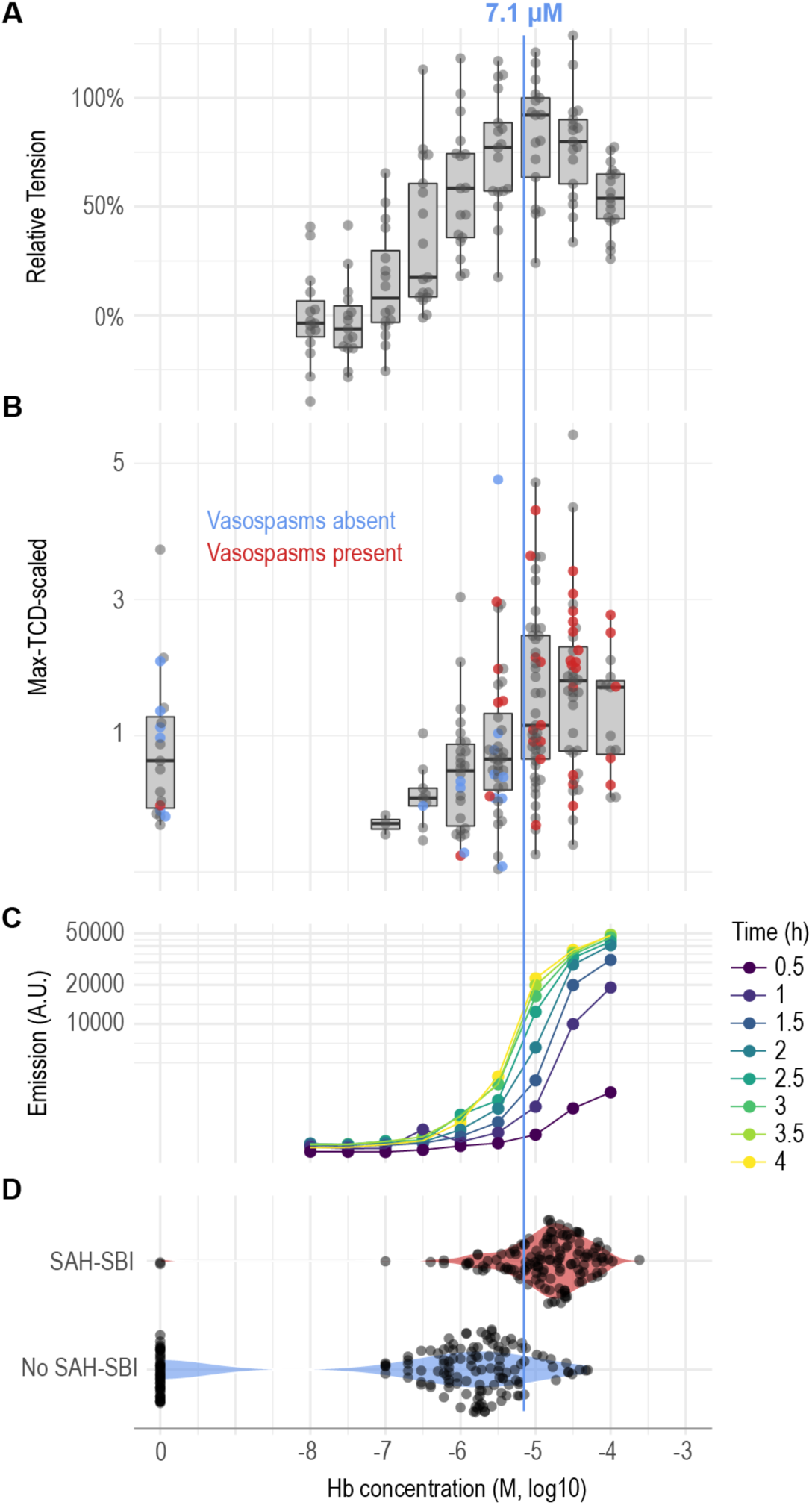
The vasoconstrictive and oxidative potential of hemoglobin. **A**. Relative tension dose-response curve for hemoglobin (Hb). **B**. Patient Transcranial Doppler (TCD) velocity (max-TCD-scaled) correlated to CSF-Hb levels. The presence of angiographic vasospasms (aVSP) is displayed as a color overlay. **C**. Formation of malondialdehyde (MDA) in response to different concentrations of Hb determined by a TBARS assay. **D**. Correlation between measured CSF-Hb and the presence of secondary brain injury (SAH-SBI). *The box in the boxplots bounds the interquartile range (IQR) divided by the median, while the whiskers extend to the highest and lowest value within the 1*.*5 x IQR, respectively*.

CSF-Hb values stratified by good and poor functional outcome at 3-month follow up based on GOSE and mRS score are shown in Figure 3D and Supplementary Figure 4H, respectively. A GAM with spline fit (4 knots) for time (day after aSAH) and a random-effect for the individual patients provided very strong evidence for a positive association of CSF-Hb to a poor functional outcome at 3-month follow up, as expressed by the GOSE (partial dependence = 0.71, SD = 0.10, p < 0.0001) and mRS (partial dependence = 0.72, SD = 0.10, p < 0.0001).

### Basilar artery vasoconstriction and lipid peroxidation occur in the concentration range of patient CSF-Hb

Our clinical data suggested CSF-Hb as an upstream mediator of SAH-SBI. To further investigate a potential pathophysiological association, we studied whether ex vivo vasoconstriction and lipid peroxidation with Hb exposure correlated with the range of CSF-Hb concentrations observed in our cohort. We established a new model of vascular function with porcine basilar arteries that were precontracted with PGF2α and dilated by an intrinsic endothelial NO synthase (eNOS) response. This setup allowed us to study the effect of Hb on endogenous NO reserve capacity. In contrast to our prior proof-of-concept studies *(16)*, which were not designed to validate the pathophysiological effects of predefined Hb concentrations, no supplemental NO donor was used. Figure 4A demonstrates the sigmoid dose-response curve, which shows a steep increase in vascular tension between 10^−6.5^ and 10^−6^ M Hb and maximum contraction near 10^−5^ M Hb.

An almost identical curve was obtained when we plotted the max-TCD-scaled values of our patient cohort against the measured CSF-Hb concentrations (Fig. 4B). The samples from those patients with aVSPs (red dots) in the majority contained CSF-Hb at concentrations of 10^−5^ M or more.

To investigate the oxidative potential associated with CSF-Hb, we measured the generation of MDA in a mixture of Hb and rLP containing unsaturated phosphatidylcholine, which is the main physiological lipid substrate for in vivo Hb peroxidation reactions *(18)*. The dose-response curves shown in Figure 4C demonstrates that the steepest increase in MDA formation also occurred with Hb at concentrations between 10^−5.5^ and 10^−5^ M with reaction incubation times ranging from 0.5 to 4 h.

Figure 4D plots the patient’s CSF-Hb concentrations stratified by the presence of SAH-SBI. This demonstrates that the optimal Youden index of CSF-Hb for SAH-SBI, which arithmetically yields the optimal ratio between sensitivity and specificity, nearly coincides with the CSF-Hb concentration at maximal basilar artery vasoconstriction, the maximum TCD values and the concentration at which the rate of TBARS generation is highest.

Collectively, the results of the vascular function and lipid peroxidation studies reveal critical inflection points for toxic Hb-effects at concentrations overlapping the CSF-Hb concentrations found in patients with SAH-SBI.

### Depletion and neutralization of CSF-Hb mitigates pathological vasoconstriction and lipid peroxidation

To mechanistically link CSF-Hb in patients with SAH-SBI, we performed Hb depletion and neutralization experiments. Porcine basilar artery segments that were immersed in CSF depleted of Hb with a haptoglobin affinity column (Fig. 5A) had a lower relative tension than segments immersed in the same CSF sample in which the original CSF-Hb concentration had been restored with highly purified Hb (Fig. 5B). This experimental setup controlled for potential nonspecific removal of vasoactive substances by the haptoglobin affinity column. Additionally, we repeated a Hb dose-response experiment to confirm the anti-vasospastic effect of soluble haptoglobin (phenotype 1-1, Hp 1-1). Across the Hb concentration range, Hp 1-1 attenuated the contractile force of basilar arteries (Fig. 5C). We also quantified the oxidative potential of CSF samples from patients after aSAH. We found variable increases in MDA after lipoprotein exposure between CSF samples that were collected during the high-risk phase (weeks 0.5-2) compared to baseline CSF samples (collected on day 1) (Fig. 5D). The addition of the Hb-scavenger Hp 1-1 at equimolar concentrations to CSF-Hb effectively attenuated MDA formation. The addition of the heme-scavenger hemopexin at an equimolar concentration to CSF-Hb attenuated MDA formation to an even greater extent. These ex vivo experiments show the effectiveness of Hb and heme scavengers in mitigating the vasoconstrictive, as well as the oxidative effects of CSF-Hb, and thereby support further evaluation for their clinical application of patients after aSAH.

**Fig. 5.**
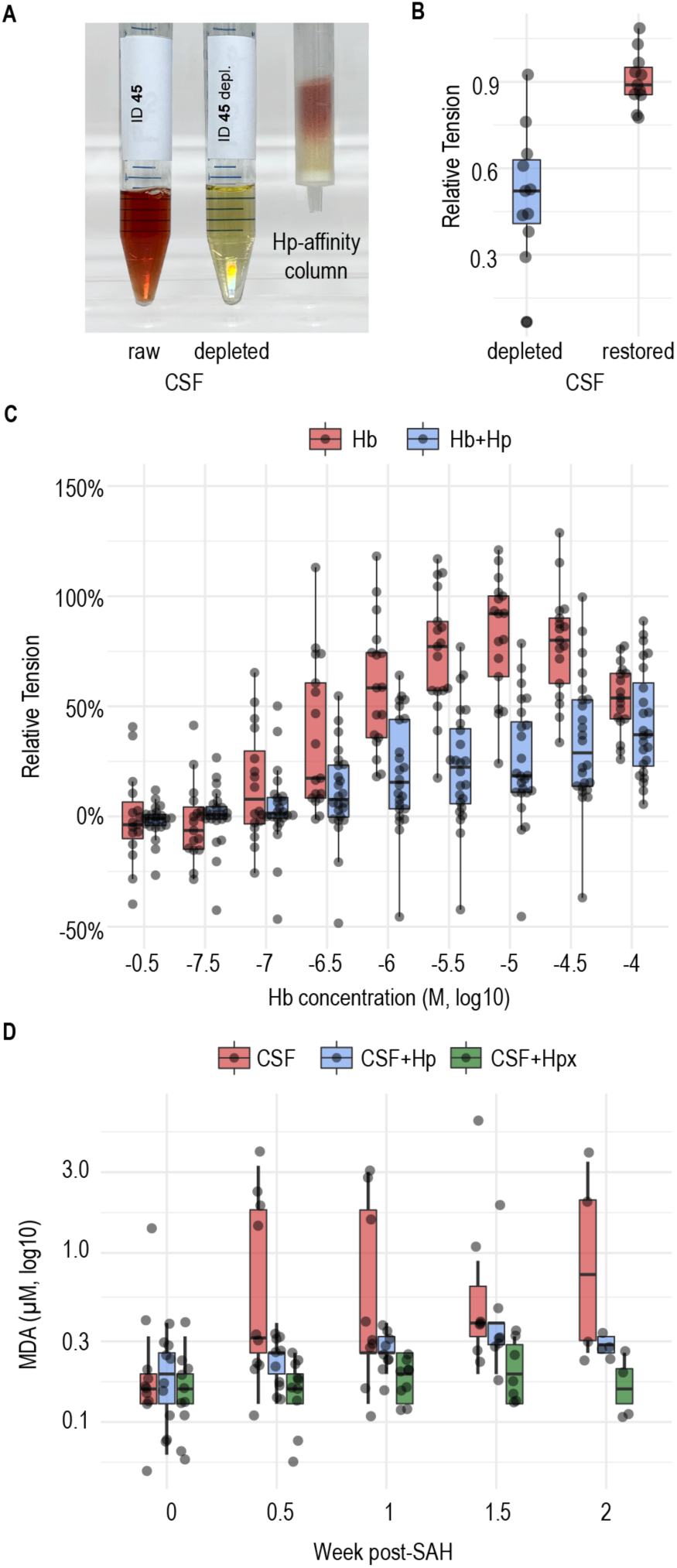
Protective effects of scavenger proteins against the vascular and oxidative effects of hemoglobin. **A**. Depletion of Hb from CSF using a haptoglobin (Hp) affinity column. **B**. Vascular tension of vessels immersed in Hb-depleted CSF and Hb-restored CSF. **C**. Dose-response curve for Hb with (data from Fig. 4A) or without Hp. **D**. The oxidative potential of patient CSF over time after aneurysmal subarachnoid hemorrhage (aSAH) with and without the addition of Hp or hemopexin (Hpx), as assessed by malondialdehyde (MDA) quantification with a thiobarbituric acid-reactive substances (TBARS) assay. *The box in the boxplots bounds the interquartile range (IQR) divided by the median, while the whiskers extend to the highest and lowest value within the 1*.*5 x IQR, respectively*.

## DISCUSSION

The main objective of this study was to investigate the clinical association of CSF-Hb with SAH-SBI and the diagnostic accuracy of daily CSF-Hb measurements for SAH-SBI. Based on a prospective cohort of 47 patients and 415 CSF samples, we provide very strong evidence for a positive association between CSF-Hb and the occurrence of SAH-SBI. In addition, CSF-Hb markedly exceeded the diagnostic accuracy of established prediction methods. Also, we aimed to rationalize the role of CSF-Hb as a targetable driver of SAH-SBI. We showed that, within a clinically relevant concentration range, CSF-Hb induced vasoconstriction and oxidized unsaturated lipids ex vivo, suggesting that it might act as an upstream toxin. The Hb-scavenger haptoglobin and the heme-scavenger hemopexin effectively counteracted both toxicity mechanisms within the clinically relevant dose range. Collectively, the strong clinical association between high CSF-Hb levels and SAH-SBI, the underlying pathophysiological rationale and the favorable effects of haptoglobin and hemopexin position CSF-Hb as a highly attractive biomarker and potential drug target.

In this study, EVD sampling-based CSF-Hb showed a diagnostic accuracy for SAH-SBI that considerably exceeded that of existing methods. While often used in clinical practice, established clinical (WFNS, Hunt & Hess) and radiological (BNI, modified Fisher) scores show a limited predictive accuracy for SAH-SBI *(19–22)*. The performance of combined scores or machine-learning-based models for predicting SAH-SBI is also only slightly better than that of the established clinical and radiological scores *(23, 24)*. The value of daily assessments of arterial flow velocity in the large cerebral arteries using TCD is controversial because of its limited diagnostic precision and high interrater variability *(25, 26)* it cannot be applied in one-fifth of patients due to anatomical reasons *(27)*. Due to the lack of superior alternatives, however, TCD is recommended by current guidelines and has been adopted by many clinical centers *(10, 28)*. In our cohort, the diagnostic accuracy of TCD for SAH-SBI was clearly below that of CSF-Hb. Moreover, literature-reported performance measures of proposed biomarkers for SAH-SBI are also substantially inferior to CSF-Hb *(29, 30)*. In addition to its high diagnostic accuracy for SAH-SBI, CSF-Hb is characterized by its simple and reliable analytics. Precise CSF-Hb values could be determined bedside with a simple two-step procedure consisting of a centrifugation step to remove cells and debris from the CSF and subsequent spectrophotometry with automated spectral deconvolution. Furthermore, spectrophotometry allows the absolute quantification of CSF-Hb without parallel measurements of calibration samples or to generate standard curves. Collectively, these advantages significantly reduce analytical costs and turnaround times and may enhance the widespread applicability and use of CSF-Hb quantification.

We and others have experimentally determined that Hb induces pathophysiological processes reflecting specific features of SAH-SBI, such as vasospasm, lipid oxidation, and neuronal damage *(16, 17, 31)*. However, a significant limitation of prior studies was that the explored Hb and heme concentrations were not based on a rational assessment of relevant CSF-Hb concentrations in patients. Indeed, our previous experiments in a sheep vasospasm model *(16)* defined effects of CSF-Hb at concentrations at least tenfold above the critical inflection point implied by our current patient data. Furthermore, our previous ex vivo vasospasm model was incapable of defining physiologically relevant vasoconstrictive Hb concentrations because we used exogenous NO donors and, therefore, artificially set the concentration threshold at which Hb interrupted vasodilatory NO signaling. To address these issues, we redesigned our ex vivo model to investigate how CSF-Hb interferes with an intrinsic vasodilator response. This was achieved by combining an algorithm-based prestretching protocol PGF2α as a pre-contracting agent, which induced endogenous, endothelial NO synthesis. With this new model, we confirmed that Hb is the major vasoconstrictor in patient CSF by comparing the vasoconstrictive effect of Hb-depleted and Hb-restored CSF. Additionally, we were able to determine that the active dynamic range of Hb concentrations in the ex vivo vasospasm model overlaps with critical inflection points that determine TCD flow velocities in large cerebral arteries and the presence or absence of SAH-SBI in our patient cohort. Within the same clinically relevant concentration range, we found that CSF-Hb was highly active as a lipid oxidant, indicating oxidative Hb toxicity to contribute to nonischemic neuronal damage. We attributed the lipid oxidation activity in patient CSF to the downstream Hb metabolite heme. Its selective neutralization with hemopexin virtually blocked the formation of MDA, while haptoglobin partially inhibited MDA formation.

Although our study followed a strongly hypothesis-driven approach, we attempted to further assess the phenomenon of CSF-Hb accumulation and toxicity within the broader context of biological processes reflected by the temporal dynamics of the CSF proteome. Based on LC-MS/MS analysis of Hb-depleted CSF from the two weeks high-risk phase for SAH-SBI, we found the strongest signals for those proteins suggestive of RBC lysis and macrophage accumulation. RBC lysis was indicated by an increase in erythrocyte glycolytic and antioxidant enzymes (e.g., CA1, CA2, CAT, and ALDOA) over time, while lineage-specific soluble cell surface receptors (e.g., CD163, CD14, CSF1R and TIMP-1) indicated the rapid and sustained accumulation of macrophages. Prior studies have suggested that leptomeningeal macrophages with enhanced expression of the Hb scavenger receptor CD163 accumulate in the subarachnoid space after aSAH *(32, 33)*. In in vitro studies and in hemolytic mice, macrophages exposed to damaged RBCs, Hb, or heme acquired an idiosyncratic phenotype supporting an adaptive response consisting of heme detoxification, iron sequestration, and inflammatory suppression, aiming at promoting hematoma resolution and wound healing *(34–37)*. The time course of the macrophage protein signatures in our patients coincided well with the early peak in CSF bilirubin, which reflects metabolic heme degradation by heme oxygenase. We found bilirubin to reach a plateau on days 3 to 4 after aSAH, followed by an increase in CSF biliverdin. This implies a saturation of the cumulative metabolic capacity of resident and recruited macrophages in the subarachnoid space at that time point (Fig. 6). Therefore, the delayed increase in CSF-Hb concentrations to toxic levels appears to be the combined result of the accelerated lysis of RBCs and the saturated capacity of erythrophagocytic and Hb-clearing macrophages. The accumulation of RBC- and macrophage-proteins occurred even with the rapid depletion of plasma proteins, such as apolipoproteins, coagulation factors and haptoglobin, reflecting rapid protein clearance from the CSF during the initial days after aSAH.

**Fig. 6.**
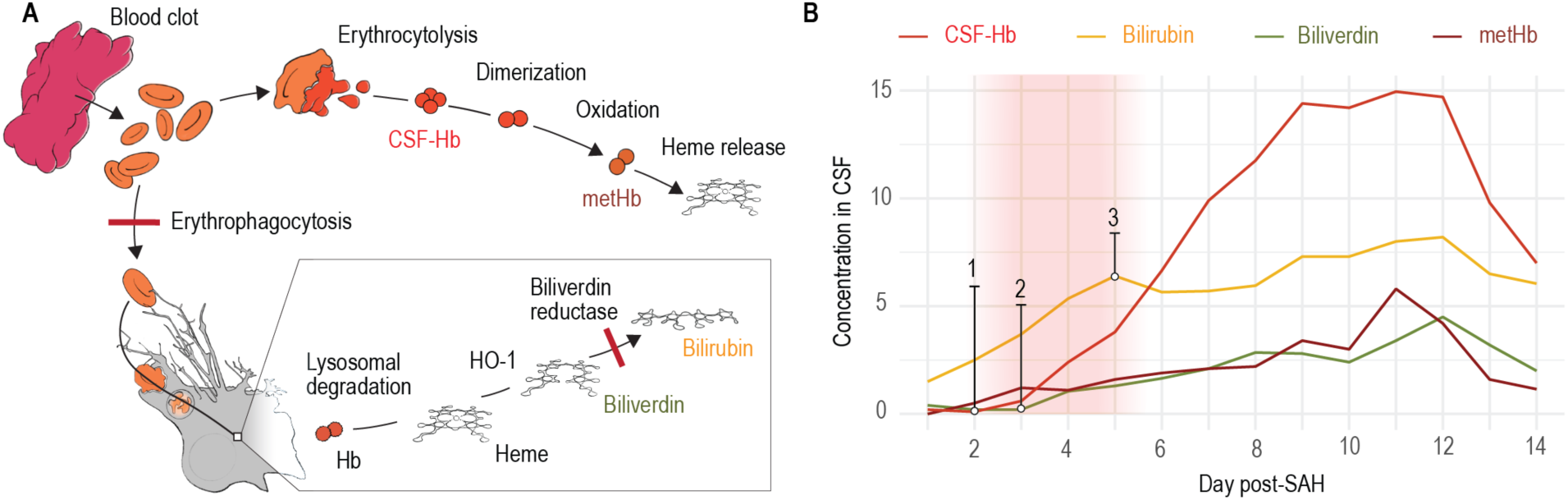
The pathophysiology of cerebrospinal fluid hemoglobin and heme metabolites after aneurysmal subarachnoid hemorrhage. **A**. Schematic illustration of the pathophysiological processes in the cerebrospinal fluid (CSF) microenvironment after aneurysmal subarachnoid hemorrhage (aSAH). Erythrophagocytosis and the consecutive intracellular processes of hemoglobin (Hb) degradation and heme metabolization are shown at the bottom. The saturation of the biliverdin reductase and the erythrophagocytosis is indicated by red bars. Erythrocytolysis in the subarachnoid CSF space is shown at the top. Liberated Hb is oxidized to methemoglobin (metHb) before heme is released. **B**. CSF concentrations of Hb (CSF-Hb), bilirubin, biliverdin and metHb over time with the assumed saturation period of biliverdin reductase and phagocytosis indicated by a red shaded area, resulting in an increase of CSF-Hb (1) and biliverdin (2) as well as the plateauing of bilirubin (3).

Our data reinforce the idea of CSF-Hb as an attractive drug target to prevent and treat SAH-SBI. This is supported by the strong clinical association between CSF-Hb and SAH-SBI, the underlying pathophysiological rationale behind this association, and the favorable functional effects of haptoglobin and hemopexin in neutralizing CSF-Hb in a clinically relevant concentration range for toxicity ex vivo. Haptoglobin is part of the physiological scavenger system for extracellular Hb, providing the most upstream antagonization of Hb toxicity by preventing vascular and tissue translocation of the large Hb-haptoglobin complex, blocking heme release, stabilizing Hb radical reactions, and accelerating Hb clearance by the macrophage CD163 receptor pathway *(18, 38–40)*. In our CSF analysis, endogenous haptoglobin was shown to be cleared from the CSF within the first few days after aSAH before the subsequent release of relevant amounts of CSF-Hb. Thus, the protective function of endogenous haptoglobin seems negligible. In a previous animal study, intraventricularly administered haptoglobin inhibited Hb delocalization from the CSF into the brain parenchyma and smooth muscle cell layer of cerebral arteries, preventing Hb-induced aVSP *(16)*. Our findings support this protective function of haptoglobin by demonstrating its anti-vasoconstrictive and antioxidant effects at a clinically relevant Hb concentration range. The antioxidant function of the heme scavenger protein hemopexin even surpassed that of haptoglobin. This suggests that a fraction of heme had already been released from oxidized or degraded CSF-Hb, forming a pool of oxidative free heme that remained associated with low-affinity heme-binding proteins. Therefore, it appears reasonable that the therapeutic combination of intraventricularly administered haptoglobin and hemopexin may exert synergistic protection.

The fact that the CSF sampling in this study was conducted via an EVD located in the lateral ventricle may represent a certain limitation regarding some of the pathophysiological conclusions. The measured CSF-Hb concentrations represent local conditions in the lateral ventricle and do not necessarily correspond to the conditions in other CSF compartments (e.g., basal cisterns). Although, patients with IVH showed a pattern with an early peak and subsequent plateau, patients without IVH reached comparable CSF-Hb levels later on. This suggests the presence of a considerable redistribution of CSF-Hb across the whole CSF space, leading to a rather homogeneous distribution over time. A potential limitation of the clinical data results from the high number of missing observations in the daily assessment of DIND, DCI or aVSP. This is intrinsic in nature, as the neurological assessment of patients with aSAH is frequently limited due to intubation and sedation, and cranial imaging is not conducted every day. The aim of using a strict definition of DIND, DCI or aVSP was to exclude uncertainties in the acquired data and to increase the reliability in the association between CSF-Hb and SAH-SBI. The homogeneous distribution of the measured CSF-Hb values within the NAs argues against a systematic error. In addition, our clinical study is limited by a selection of severe clinical cases due to the inherent need for an EVD and its unicentric nature. As a next step, a multicenter validation study is needed to generalize our findings across different clinical centers and to determine a suitable CSF-Hb cutoff.

## MATERIALS AND METHODS

### General

This study was approved by the local ethical review board (KEK ZH 2016-00439). Written informed consent was obtained from all patients or their legal representatives. All experiments conformed with the appropriate biosafety regulations and were carried out under appropriate working conditions. The results are reported in accordance with the STROBE statement *(41)*.

### Study population

Clinical data and CSF samples were obtained from a prospective consecutive cohort of aSAH patients admitted to the Neurointensive Care Unit of the Department of Neurosurgery, University Hospital Zurich over a 3-year period (April 2017 to March 2020). The eligibility criteria were as follows: (i) age > 18 years; (ii) radiological diagnosis of subarachnoid hemorrhage; (iii) exclusion of non-aneurysmal subarachnoid hemorrhage (e.g., trauma, perimesencephalic subarachnoid hemorrhage); and (iv) insertion of an external ventricular drain (EVD) as part of the standard of care.

### Clinical data acquisition

Patient data collection was performed by the treating physician blinded to the CSF measurements. The following baseline features of the patients were assessed at the time of diagnosis: demographic characteristics (age, sex), clinical scores (World Federation of Neurosurgical Societies [WFNS] grade, Hunt & Hess grade, initial Glasgow Coma Scale [GCS]) and radiological parameters (aneurysm location, aneurysm size [maximal aneurysm diameter], hematoma volume [see below], presence of intraventricular hemorrhage [IVH], modified Fisher grading, and Barrow Neurological Institute [BNI] grading) (Supplementary Fig. 1A). Clinical data was collected prospectively during the 14-day high-risk phase for SAH-SBI based on standardized monitoring at the Neurointensive Care Unit and as part of the 3-month follow-up. General patient management conformed to current guidelines of the Neurocritical Care Society and the American Heart Association *(10, 11)*. The presence of DINDs, DCI and aVSPs was assessed on a daily basis during the 14-day sampling period. DIND was defined as a new focal neurological deficit or a decrease in GCS score of at least 2 points for at least 2 hours that was not attributable to any other identifiable cause, such as hydrocephalus, epileptic seizure or systemic factors. In case the patient’s neurocognitive state could not be clinically assessed (e.g., due to sedation), the clinical data was denoted as not available (NA). DCI was defined as new ischemia or new infarction on MRI or CT that was not attributable to surgical intervention. An aVSP was defined as a new narrowing of a cerebral artery based on digital subtraction angiography, CT angiography or MR angiography. When an imaging procedure was not carried out on a given day, the findings were denoted as NA. Thus, the strict categorization as NA served to prevent speculative clinical assessment regarding DIND, DCI and aVSP. SAH-SBI was defined as the composite outcome of DIND, DCI and aVSP. In addition, flow velocity in the bilateral middle, anterior and posterior cerebral arteries was assessed daily using Transcranial Doppler (TCD) sonography. Complications (meningoencephalitis, cerebral salt wasting, diabetes insipidus, pneumonia, Takotsubo cardiomyopathy, nonconvulsive status epilepticus) were assessed daily during the 14-day sampling period. Chronic hydrocephalus (ventriculoperitoneal shunt dependency) and functional status (Glasgow Outcome Scale-Extended [GOSE] and modified Rankin Scale [mRS]) were evaluated at the 3-months follow-up.

### Subarachnoid hematoma segmentation and volumetric analysis

Based on the initial CT scan, the volume of the subarachnoid hematoma was manually delineated using 3DSlicer 4.11.0 *(42)*. The preprocessing prior to delineation consisted of brain extraction whereby the largest cavity from a 200 HU thresholded skull mask was extracted *(43)*. Subsequently, the brain mask was multiplied with the original CT scan and median filtered with a 5×5×3 kernel (Supplementary Fig. 1B). The volume of the hemorrhage was calculated by multiplying the voxel size by the number of segmented voxels.

### CSF sampling

CSF samples were collected from the EVD by investigators blinded to the clinical data. The decision regarding the insertion of an EVD was made independently of this study and according to the clinical standard of care. 1ml of CSF was sampled on a daily basis at approximately the same time (morning). A sampling duration from day 1 (the day after aSAH) to day 14 was used. If the patient’s EVD was removed prior to day 14, the CSF sampling was discontinued. Immediately after sample collection, the CSF was centrifuged (Capricorn CEP 2000 Benchtop centrifuge, Capricorn labs, UK) at 1500 ×g for 15 minutes, and the supernatant was stored at −80°C for further analysis (Supplementary Fig. 1C).

### Spectrophotometry of CSF-Hb and heme metabolites

Absorption spectra in the visual range between 350 and 650 nm of all CSF samples were measured on a Shimadzu UV-1800 spectrophotometer (Shimadzu, Japan). Quantification of different Hb species and their metabolites in the CSF (oxyHb [hereafter referred to as CSF-Hb], metHb, bilirubin, biliverdin) was performed using spectral deconvolution. Therefore, extinction curves for the individual substances with known concentrations were fitted to the extinction curve of CSF using a nonnegative least squares algorithm as described previously *(18)*.

### LC-MS/MS CSF proteomic analysis

Using LC-MS/MS based label-free quantification, the CSF proteomes of 18 patients were obtained at five sequential time points during the observation period (0, 0.5, 1, 1.5 and 2 weeks after aSAH) (Supplementary Fig. 1C). The samples were processed in two batches to improve generalizability, and the data was combined for analysis after normalization (85 samples in total; 48 in batch 1, 37 in batch 2). Hb, which is abundantly present in CSF following erythrolysis, was selectively removed prior to the analysis using a haptoglobin affinity column to avoid ion suppression and increase the sensitivity as previously described *(16)*. Following digestion of the Hb-depleted CSF samples with trypsin, mass spectrometry analysis was performed on a Q Exactive HF mass spectrometer (Thermo Scientific) equipped with a Digital PicoView source (New Objective) and coupled to an M-Class UPLC (Waters). Details of the proteomics workflow and sample preparation are included in the Supplementary Methods. The individual protein intensities were normalized to the mean intensity of the respective protein at baseline (week 0) and log2 transformed. The overall fold change in protein abundance was calculated as the sum of the log-ratios for each protein over the whole time course. Then, normalized protein intensities at each time point were compared to the baseline intensities (week 0) using a Wilcoxon rank-sum test, and p-values were combined over all time points using the metap package *(44)*. Temporal changes in the CSF proteome were clustered using k-means analysis, whereby the optimal number of clusters was determined visually with an elbow diagram (Supplementary Fig. 1D) using the factoextra package *(45)*.

### Neurovascular function

The neurovascular function assay (Supplementary Fig. 1E) was performed using fresh porcine basilar arteries obtained from a local abattoir (n = 12) *(16)*. The sequence of the experiment is given in Supplementary Fig. 1F. The vessels were pre-stretched to reach the optimal passive pretension (IC1 with factor k = 0.80) as previously published *(46)*. Then, 10 µM prostaglandin F2α (PGF2α; Sigma, Buchs, Switzerland) was used as a pre-contracting agent. This was followed by an endogenous NO-dependent dilation of the vessels. Subsequently, the specific experiments were performed as described below. At the end of the experiment, to determine the maximum possible tension in the absence of endogenous NO, L-N5-(1-iminoethyl)ornithine hydrochloride (L-NIO; Sigma-Aldrich, Saint Louis, US, MO) at a concentration of 10 µM was added. The reported relative tension represents the absolute tension normalized to the individual NO reserve capacity of the respective vessel, i.e., the baseline tension after the addition of PGF2α (0%) and the tension after the addition of L-NIO at the end of the experiment (100%). Vessels with a continuous tension loss throughout the experiment, resulting in a PGF2α^max^/L-NIO^max^ ratio below 0.7, were considered to be impaired and therefore excluded from the analysis.

An initial series of experiments was conducted to evaluate the vasoconstrictive potential of patient CSF during the high-risk phase for SAH-SBI and to assess the specific role of Hb in this process. For this purpose, patient CSF collected between days 3 and 14 after aSAH was selectively depleted of CSF-Hb using a haptoglobin affinity column as previously described *(16)*. In the initial phase of the experiment, the vessels were immersed in Hb-depleted CSF. Subsequently, the precise amount of Hb that had been removed from the CSF was restored, in order to determine the specific impact of CSF-Hb on vascular tension. In a second series of experiments, the influence of increasing Hb exposure on vascular tension was assessed. For that purpose, the vessels were immersed in Krebs-Henseleit buffer and exposed to increasing concentrations of Hb (10^−4^ M to 10^−8^ M Hb in half log10 steps). To evaluate the effect of the hemoglobin-scavenger haptoglobin, a third series of experiments was performed with identical Hb concentrations, but an equimolar amount of haptoglobin was added.

### Hb, haptoglobin, hemopexin and reconstituted lipoprotein (rLP)

Hb for use in ex vivo experiments was purified from expired human blood concentrates as previously described *(47)*. Hb concentrations were determined by spectral deconvolution as described above and are given as molar concentrations of total heme (1 M Hb tetramer is equivalent to 4 M heme). For all Hb used in these studies, the fraction of ferrous Hb (HbFe^2+^O2) was always greater than 98%, as determined by spectrophotometry. Purified haptoglobin (phenotype 1-1) and hemopexin from human plasma, as well as reconstituted lipoprotein (rLP) were obtained from CSL Behring, Bern, Switzerland.

### Thiobarbituric acid-reactive substances (TBARS) assay

The oxidative potential of the CSF samples was quantified by measuring the formation of malondialdehyde (MDA), the final product of lipid peroxidation, after incubation with rLP (Supplementary Fig. 1G). Using a 96-well plate, 25 µL of CSF was mixed with 5 µL of rLP (3 mg/mL) and incubated at 37°C for 24 hours. Subsequently, the concentration of MDA was measured using a TBARS assay *(18)*. In short, 125 µL of 750 mM trichloroacetic acid in 1 M HCl was added to the samples, followed by vortexing (5 seconds) and the subsequent addition of 100 µL of 25 mM 2-thiobarbituric acid in 1 M NaOH. After an incubation period of 60 minutes at 80°C, the TBARS in the supernatant was quantified. For the absolute quantification of MDA, the absorption at 600 nm was subtracted from the absorption at 532 nm, and the concentration was calculated with a molar extinction coefficient of 0.156 mM^−1^cm^−1^. To achieve more sensitive but relative quantification, the fluorescence emission was measured at 550 nm with 510 nm used as the excitation wavelength.

### Statistical analyses

Statistical analyses were performed using R 3.6.3 *(48)*. The detailed statistical methods as well as the versions of the used R-packages are given in Supplementary Methods. Descriptive statistics are presented as absolute numbers (n) and proportions (%) for categorical variables, whereas continuous variables are shown as mean and standard deviation (SD). Due to the exploratory nature of the study, no level of statistical significance was defined; instead, the results were interpreted based on the level of evidence for an association as follows: p < 0.001: very strong evidence; p < 0.01: strong evidence; p < 0.05 evidence; p < 0.1 weak evidence; and p > 0.1: no evidence *(49)*.

## Data Availability

The authors confirm that the data supporting the findings of this study are available within the article and its Supplementary Materials. In addition, all anonymized datasets used in this study along with the statistical code are provided in the Supplementary Materials (for an overview, see Supplementary Table 3). The use of dynamic reporting guarantees full reproducibility of the results given data and code.

## Supplementary Materials

Materials and Methods. LC-MSMS CSF proteome analysis.

Materials and Methods. Statistical analyses.

Fig. S1. Methods.

Fig. S2. Individual profiles of cerebrospinal fluid hemoglobin and heme metabolites.

Fig. S3. Temporal profiles of cerebrospinal fluid hemoglobin stratified by angiographic vasospasms, delayed cerebral ischemia and delayed ischemic neurological deficits.

Fig. S4. Association between cerebrospinal fluid hemoglobin with secondary brain injury after aneurysmal subarachnoid hemorrhage and modified Rankin Scale.

Table S1. Temporal profiles of cerebrospinal fluid hemoglobin and heme metabolites. Table S2. Cohort outcome characteristics.

Table S3. Datasets.

Data file S1. Proteomics (raw intensities).

Data file S2. Proteomics (normalized intensities).

Data file S3. Photospectrometric CSF measurements.

Data file S4. Clinical patient data.

Data file S5. TBARS from patient CSF (batch 1).

Data file S6. TBARS from patient CSF (batch 2).

Data file S7. TBARS Hb dose range.

Data file S8. Vascular function Hb dose range.

Data file S9. Vascular function Hb + Hp dose range.

Data file S10. Vascular function normalization plot.

Data file S11. Vascular function with patient CSF.

Statistical code S1.

## Acknowledgments

We thank Nadja Schulthess and Kerstin Hansen for technical support. We thank the staff of the Functional Genomic Center Zurich (FGCZ) for support with the mass spectrometry analysis of CSF samples.

## Author contributions

Conceptualization: KA, RMB, CAS, MH, DJS.

Methodology: KA, RMB, CAS, BRT, UH, JWD, MH, DJS.

Investigation: KA, RMB, CAS, BRT, SW, JW, MS, UH, EK, MH, DJS.

Visualization: RH.

Funding acquisition: RMB, MH, DJS.

Project administration: LR, EK, MH, DJS.

Supervision: MH, DJS.

Writing – original draft: KA, RMB, MH, DJS.

Writing – review & editing: KA, RMB, CAS, BRT, FV, SW, JW, MS, UH, JWD, RH, LR, EK, MH, DJS.

