## Supplementary Material for "Cerebrospinal fluid hemoglobin drives subarachnoid hemorrhage-related secondary brain injury"

**LC-MSMS CSF proteome analysis**

*Sample preparation*

Total protein concentration was estimated using the Qubit® Protein Assay Kit (Life Technologies, Zurich, Switzerland). The samples were then prepared by using a commercial iST Kit (PreOmics Phoenix, Germany) with an updated version of the protocol. Briefly, 20 µg of protein was solubilized in 'Lyse' buffer, boiled at 95 °C for 10 minutes and processed with HIFU for 30 s, setting the ultrasonic amplitude to 85%. Afterwards, the samples were transferred to the cartridge and digested by adding 50 µl of the 'Digest' solution. After 60 minutes of incubation at 37 °C, the digestion was stopped with 100 µl of Stop solution. The solutions in the cartridge were removed by centrifugation at 3800 g, while the peptides were retained by the iST-filter. Finally, the peptides were washed, eluted, dried and resolubilized in 20 µL of buffer (3% acetonitrile, 0.1% formic acid) buffer for LC-MS analysis.

*Liquid chromatography-mass spectrometry analysis*

Mass spectrometry analysis was performed on an Orbitrap Fusion Lumos (Thermo Scientific) equipped with a Digital PicoView source (New Objective) and coupled to a M-Class UPLC (Waters). Solvent composition at the two channels was 0.1% formic acid for channel A and 0.1% formic acid, 99.9% acetonitrile for channel B. For each sample 1 µL of diluted peptides were loaded on a commercial MZ Symmetry C18 Trap Column (100Å, 5 µm, 180 µm x 20 mm, Waters) followed by nanoEase MZ C18 HSS T3 Column (100Å, 1.8 µm, 75 µm x 250 mm, Waters). The peptides were eluted at a flow rate of 300 nL/min by a gradient from 5 to 22% B in 77 min, 32% B in 10 min and 95% B for 10 min. Samples were acquired in a randomized order. The mass spectrometer was operated in data-dependent mode (DDA) acquiring a full-scan MS spectra (300−1'500 m/z) at a resolution of 120'000 at 200 m/z after accumulation to a target value of 500'000. Data-dependent MS/MS were recorded in the linear ion trap using quadrupole isolation with a window of 0.8 Da and HCD fragmentation with 35% fragmentation energy. The ion trap was operated in rapid scan mode with a target value of 10'000 and a maximum injection time of 50 ms. Only precursors with intensity above 5'000 were selected for MS/MS and the maximum cycle time was set to 3 s. Charge state screening was enabled. Singly, unassigned, and charge states higher than seven were rejected. Precursor masses previously selected for MS/MS measurement were excluded from further selection for 20 s, and the exclusion window was set at 10 ppm. The samples were acquired using internal lock mass calibration on m/z 371.1012 and 445.1200. The mass spectrometry proteomics data were handled using the local laboratory information management system (LIMS)(Türker *et al.*, 2010).

*Protein identification and label free protein quantification*

The acquired raw MS data were processed by MaxQuant (version 1.6.2.3), followed by protein identification using the integrated Andromeda search engine(Cox and Mann, 2008). Spectra were searched against a Swissprot Homo sapiens reference proteome (taxonomy 9606, version from 2019-07-09), concatenated to its reversed decoyed fasta database and common protein contaminants. Carbamidomethylation of cysteine was set as fixed modification, while methionine oxidation and N-terminal protein acetylation were set as variables. Enzyme specificity was set to trypsin/P allowing a minimal peptide length of 7 amino acids and a maximum of two missed-cleavages. MaxQuant Orbitrap default search settings were used. The maximum false discovery rate (FDR) was set to 0.01 for peptides and 0.05 for proteins. Label free quantification was enabled and a 2 minutes window for match between runs was applied. In the MaxQuant experimental design template, each file is kept separate in the experimental design to obtain individual quantitative values. The MaxQuant results were loaded into Scaffold (Proteome Software Inc.) to validate the peptide and protein identifications. Only proteins identified with at least 2 peptides were considered for follow up analysis.

**Statistical analyses**

*Statistical methods*

The linear mixed model analysis was performed with the lme4 package (Bates *et al.*, 2015). For gene set enrichment analysis (GSEA), we used GSEA software from the Broad Institute (Subramanian *et al.*, 2005) and the hallmark gene sets of the Molecular Signature Database (MSigDB) (Liberzon *et al.*, 2015). As input, we created a ranked list of the identified proteins in patient CSF, whereby the rank score was calculated as the product of -log10 (p-value) and the sum (log-ratio). The associations between baseline measures and CSF-Hb, CSF-Hb and SAH-SBI, as well as CSF-Hb and functional outcomes were analyzed using generalized additive models (GAMs) with non-linear spline-fit for the time (days after aSAH) and a random-effect for individual patients using the mgcv package (Wood, 2017). The diagnostic accuracy of CSF-Hb for SAH-SBI, aVSP, DCI and DIND was analyzed using receiver operating characteristic (ROC) curves (ROCR package (Sing *et al.*, 2005), plotROC package (Sachs, 2017)), the corresponding area under the curve (AUC) (pROC package (Robin *et al.*, 2011)) and the optimal Youden index (ROCit package (Md Riaz Ahmed, 2019)). TCD flow velocities were first used categorically according to clinical assessment (pathological or non-pathological), which served to determine its sensitivity and specificity for aVSP. Second, as TCD-scaled, the flow velocities were used as a continuous measurement for a pathophysiological association with the measured CSF-Hb concentration, with each value being scaled to the mean flow in the corresponding vessel (anterior, middle or posterior cerebral artery) of the cohort. The max-TCD-scaled represented the maximum scaled value measured daily.

*Version of R-packages*

- metap (v1.4)(Dewey, 2020)
- factoextra (v1.0.7)(Kassambara and Mundt, 2020)
- lme4 (v1.1-23)(Bates *et al.*, 2015)

- mgcv (v1.8-31)(Wood, 2017)
- ROCR (v1.0-11)(Sing *et al.*, 2005)
- plotROC (v2.2.1)(Sachs, 2017)
- pROC (v1.16.2)(Robin *et al.*, 2011)
- ROCit (v1.1.1)(Md Riaz Ahmed, 2019)

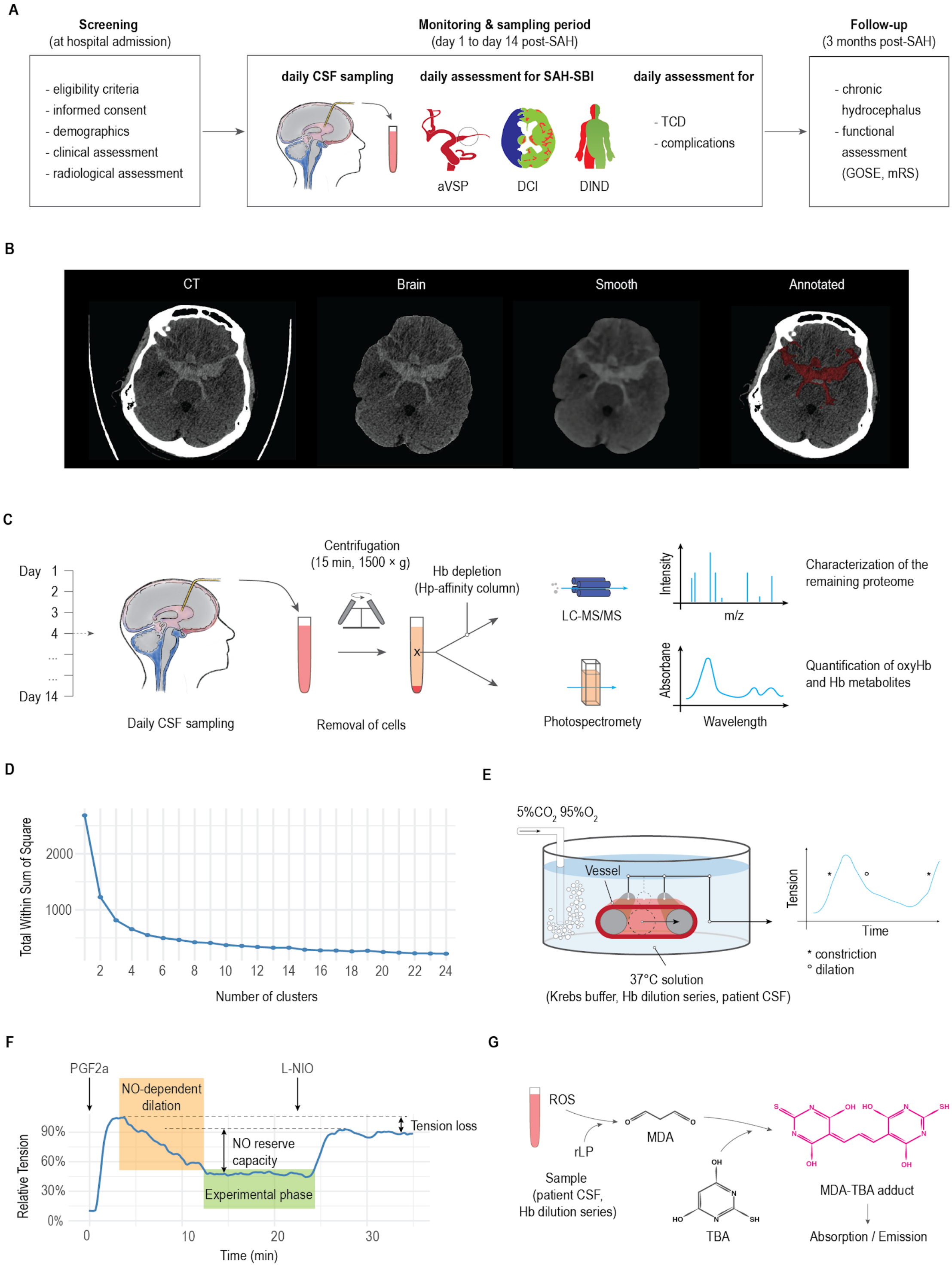


**Supplementary Fig. 1. Methods**

**A.** Flow diagram of the clinical observational study. **B.** Steps of subarachnoid hematoma segmentation for volumetric analysis. **C.** Schematic representation of daily cerebrospinal fluid (CSF) sampling via an external ventricular drain (EVD), centrifugation of the CSF and proteomic analysis of the supernatant using spectrophotometry and LC-MS/MS. **D.** Elbow plot to determine the optimal number of protein clusters. **E.** Schematic representation of the experimental setup for the neurovascular function assay. **F.** Normalization of the recorded vessel tension to the individual nitric oxide (NO) reserve capacity of the respective vessel. **G.** Schematic representation of the steps in the thiobarbituric acid-reactive substances (TBARS) assay.

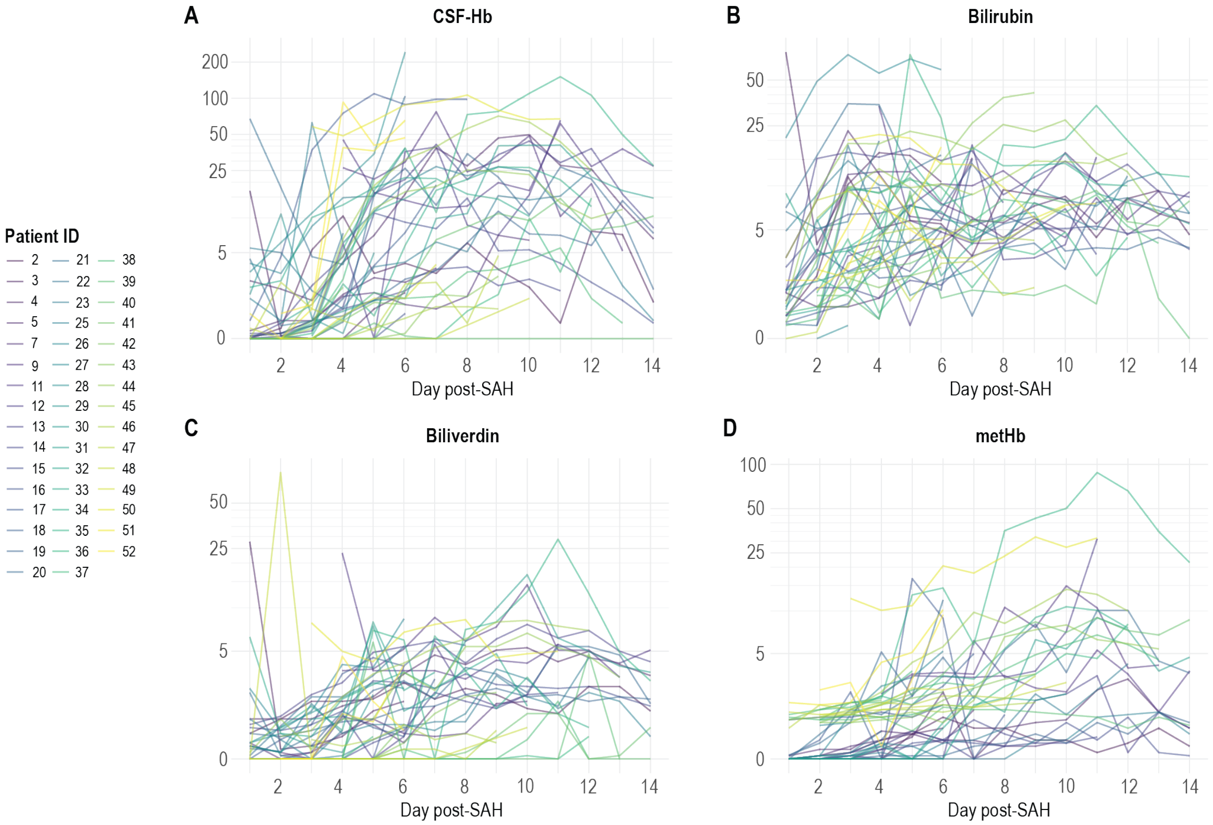


**Supplementary Fig. 2. Individual profiles of cerebrospinal fluid hemoglobin and heme metabolites**

Individual temporal profiles of (**A.**) cerebrospinal fluid hemoglobin (CSF-Hb), (**B.**) bilirubin, (**C.**) biliverdin and (**D.**) methemoglobin (metHb).

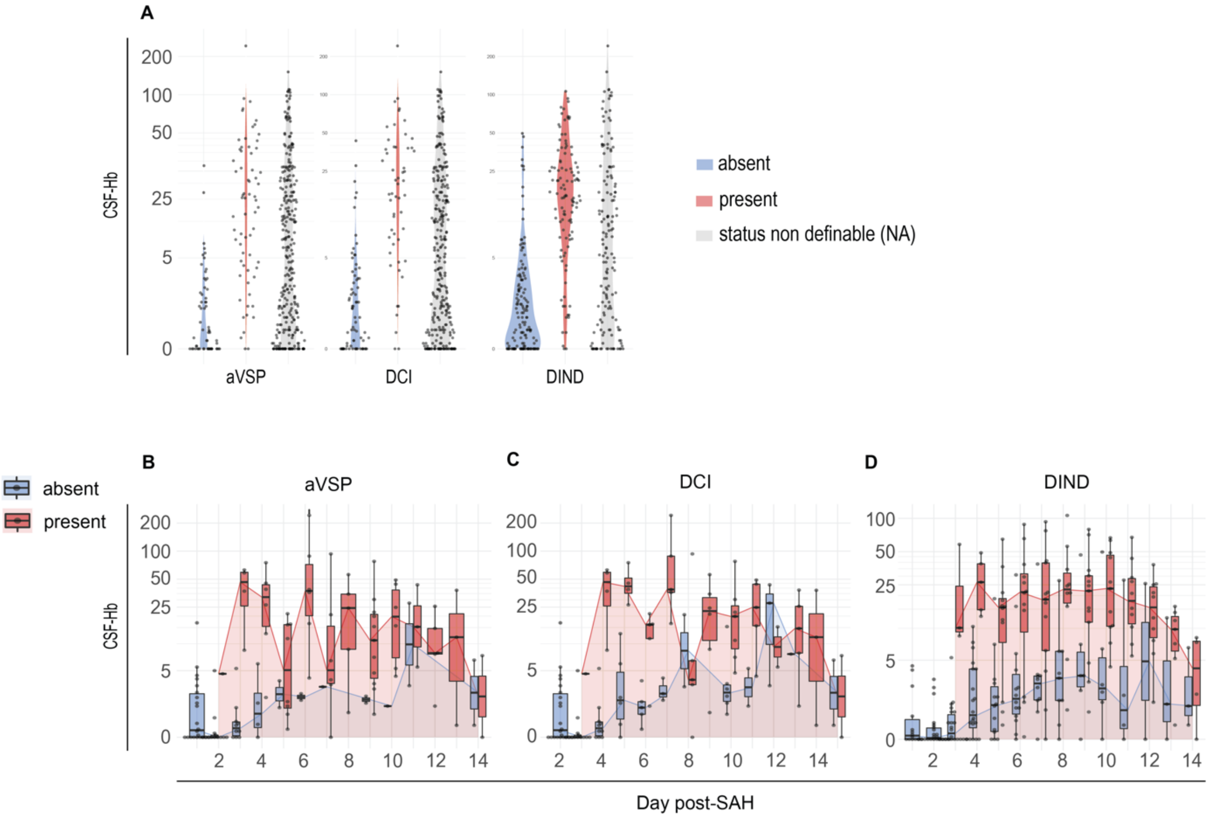


**Supplementary Fig. 3. Cerebrospinal fluid hemoglobin stratified by occurrence of angiographic vasospasms, delayed cerebral ischemia and delayed ischemic neurological deficits.**

**A.** Cerebrospinal fluid hemoglobin (CSF-Hb) in patients after aneurysmal subarachnoid hemorrhage (aSAH) stratified by presence, absence, or non-definable status of angiographic vasospasm (aVSP), delayed cerebral ischemia (DCI), and delayed ischemic neurological deficit (DIND). The temporal course of cerebrospinal fluid hemoglobin (CSF-Hb) stratified by (**B.**) angiographic vasospasms (aVSP), (**C.**) delayed cerebral ischemia (DCI) and (**D.**) delayed ischemic neurological deficits (DIND). *The box in the boxplots bounds the interquartile range (IQR) divided by the median, while the whiskers extend to the highest and lowest value within the 1.5 x IQR, respectively.*

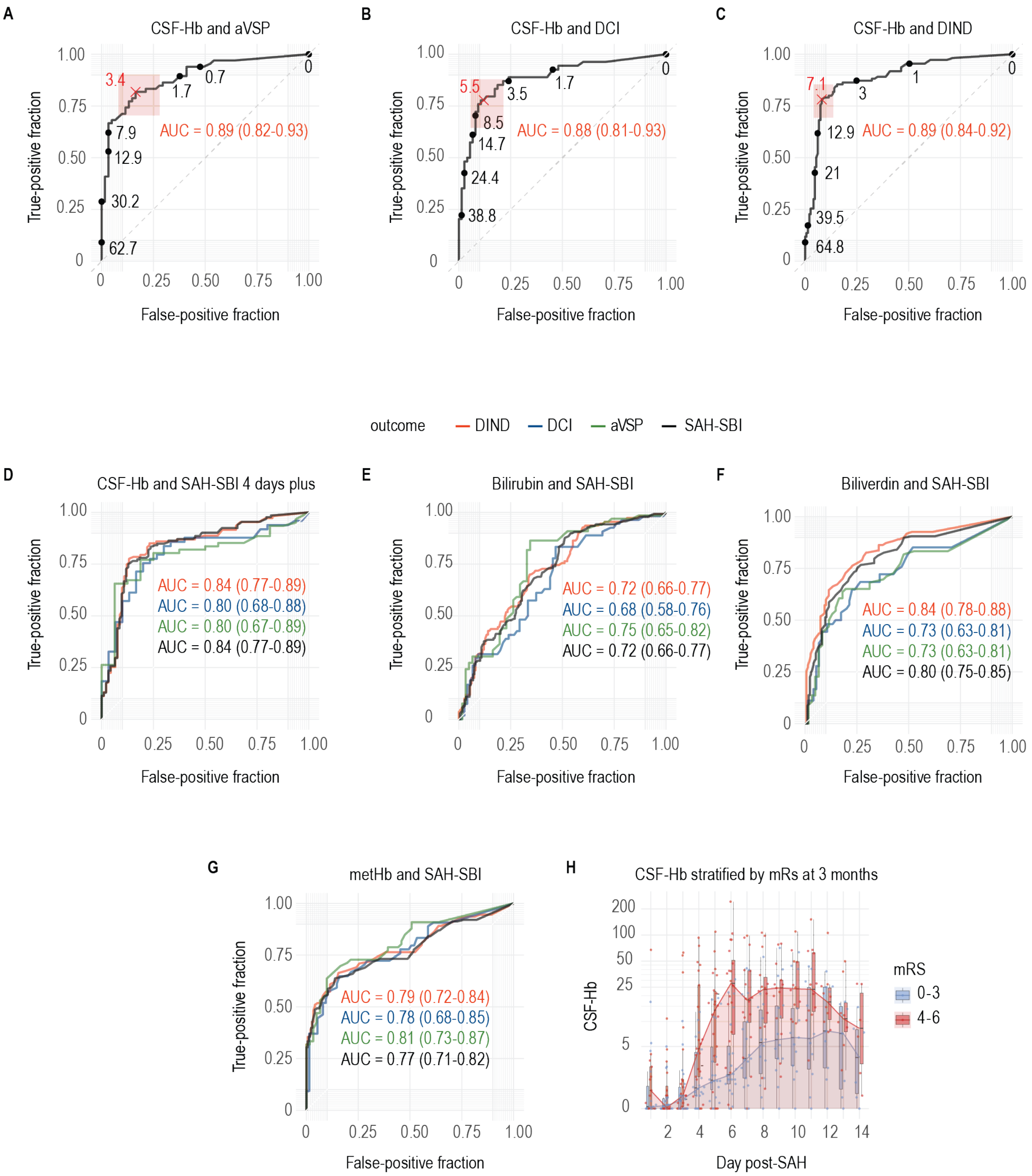


**Supplementary Fig. 4. Association between cerebrospinal fluid hemoglobin with secondary brain injury after aneurysmal subarachnoid hemorrhage and modified Rankin Scale**

**A-C.** Receiver operating characteristic (ROC) curves and areas under the curve (AUCs) of cerebrospinal fluid hemoglobin (CSF-Hb) for (**A.**) angiographic vasospasm (aVSP), (**B.**) delayed cerebral ischemia (DCI) and (**C.**) delayed ischemic neurological deficit (DIND), with the corresponding CSF-Hb measurements. The optimal Youden index and its 95% confidence interval (CI) are indicated in red. **D.** ROC curves and AUCs of CSF-Hb for aVSP, DCI, DINDs and subarachnoid hemorrhage related secondary brain injury (SAH-SBI) with data limited to the SAH-SBI high-risk period (days 4-14). **E-G.** ROC curves and AUCs of (**E.**) CSF bilirubin, (**F.**) CSF biliverdin and (**G.**) CSF methemoglobin (metHb) for aVSP, DCI, DIND and SAH-SBI. **H.** The temporal course of CSF-Hb stratified by modified Rankin Scale (mRS) at 3-month follow-up. *The box in the boxplots bounds the interquartile range (IQR) divided by the median, while the whiskers extend to the highest and lowest value within the 1.5 x IQR, respectively.*

**Supplementary Table 1. Temporal profiles of cerebrospinal fluid hemoglobin and heme metabolites**

Median, mean and 95% confidence intervals (CI) for cerebrospinal fluid hemoglobin (CSF-Hb), bilirubin, biliverdin and methemoglobin (metHb) are provided, stratified by the number of days after aneurysmal subarachnoid hemorrhage (day post-aSAH).

| Day post-SAH | CSF-Hb (µM) | | Bilirubin (µM) | | Biliverdin (µM) | | metHb (µM) | |
|---|---|---|---|---|---|---|---|---|
| | median | mean (95% CI) | median | mean (95% CI) | median | mean (95% CI) | median | mean (95% CI) |
| **1** | 0.2 | 3.9 (0-8.72) | 1.5 | 5.29 (0-10.72) | 0.4 | 1.71 (0-3.67) | 0 | 0.4 (0.15-0.65) |
| **2** | 0.1 | 0.99 (0.29-1.69) | 2.5 | 4.61 (1.9-7.32) | 0.2 | 2.53 (0-6.9) | 0.5 | 0.76 (0.5-1.02) |
| **3** | 0.6 | 4.82 (0.59-9.05) | 3.7 | 8.88 (5.04-12.72) | 0.2 | 0.7 (0.29-1.11) | 1.21 | 1.27 (0.68-1.85) |
| **4** | 2.4 | 10.5 (4.13-16.87) | 5.35 | 9.46 (6.09-12.83) | 1.05 | 1.78 (0.64-2.92) | 1.1 | 1.45 (0.89-2.01) |
| **5** | 3.8 | 12.9 (6.67-19.13) | 6.4 | 10.4 (5.92-14.89) | 1.3 | 2.13 (1.47-2.8) | 1.6 | 2.41 (1.37-3.46) |
| **6** | 6.65 | 24.99 (10.89-39.08) | 5.65 | 8.76 (5.57-11.94) | 1.65 | 2.31 (1.63-3) | 1.9 | 3.44 (2.03-4.84) |
| **7** | 9.9 | 19.52 (10.35-28.68) | 5.7 | 8.06 (5.82-10.3) | 2.1 | 2.6 (1.74-3.47) | 2.1 | 2.91 (1.68-4.15) |
| **8** | 11.75 | 21.85 (10.87-32.83) | 5.95 | 8.46 (5.41-11.5) | 2.85 | 3.06 (2.15-3.97) | 2.2 | 4.83 (1.87-7.79) |
| **9** | 14.4 | 21.89 (12.66-31.11) | 7.3 | 9.28 (6.09-12.47) | 2.8 | 3.45 (2.38-4.52) | 3.4 | 6.27 (2.46-10.08) |
| **10** | 14.2 | 24.59 (13.6-35.58) | 7.3 | 8.98 (6.38-11.58) | 2.4 | 4.39 (2.56-6.22) | 3 | 7.12 (2.63-11.6) |
| **11** | 14.95 | 28.3 (12.94-43.65) | 8 | 9.09 (6.03-12.14) | 3.4 | 4.65 (2.07-7.23) | 5.8 | 11.35 (2.86-19.83) |
| **12** | 14.7 | 20.01 (8.81-31.2) | 8.2 | 8.95 (6.82-11.07) | 4.5 | 4.05 (2.62-5.48) | 4.2 | 7.81 (0.85-14.76) |
| **13** | 9.8 | 12.95 (5.27-20.62) | 6.5 | 7.01 (5.36-8.66) | 3.2 | 2.81 (1.86-3.77) | 1.6 | 4.96 (0.26-9.66) |
| **14** | 7 | 8.91 (2.74-15.07) | 6.04 | 6.07 (4.17-7.98) | 2 | 2.35 (1.4-3.3) | 1.15 | 3.98 (0.14-7.82) |

**Supplementary Table 2. Cohort outcome characteristics**

The various manifestations of subarachnoid hemorrhage-related secondary brain injury (SAH-SBI) as well as complications and death after aSAH are shown. For manifestations of SAH-SBI, patient-wise and day-wise assessments are shown. *DCI = delayed cerebral ischemia; DIND = delayed ischemic neurological deficit; GOSE = Glasgow Outcome Scale-Extended; ICV-rtPA = intracerebroventricular recombinant tissue plasminogen activator; mRS = modified Rankin Scale; NCSE = nonconvulsive status epilepticus; rtPA = recombinant tissue plasminogen activator; aVSP = angiographic vasospasm.*

| | **Per-patient (%)** | | **Per-day (%)** | |
|---|---|---|---|---|
| **Total n** | 47 patients | | 415 samples | |
| **ICV-rtPA** | 5 (10.6) | | - | |
| **Spasmolysis** | 7 (14.9) | | - | |
| **Meningoencephalitis** | 1 (2.1) | | - | |
| **Cerebral salt wasting** | 19 (40.4) | | - | |
| **Diabetes insipidus** | 9 (19.1) | | - | |
| **Pneumonia** | 34 (72.3) | | - | |
| **Takotsubo cardiomyopathy** | 6 (12.8) | | - | |
| **NCSE** | 4 (8.5) | | - | |
| **Chronic hydrocephalus** | 24 (51.1) | | - | |
| **Death** | 14 (29.8) | | - | |
| | | *absent* | *present* | *NA* |
| **aVSP** | 23 (48.9) | 61 (14.7) | 66 (15.9) | 288 (69.4) |
| **DCI** | 21 (44.7) | 75 (18.1) | 54 (13.0) | 286 (68.9) |
| **DIND** | 30 (63.8) | 149 (35.9) | 110 (26.5) | 156 (37.6) |
| **SAH-SBI** | 31 (66.0) | 174 (41.9) | 138 (33.3) | 103 (24.8) |
| **GOSE (at 3 months)** | | | | |
| 1 | 14 (29.8) | | - | |
| 2 | 1 (2.1) | | - | |
| 3 | 10 (21.3) | | - | |
| 4 | 4 (8.5) | | - | |
| 5 | 7 (14.9 | | - | |
| 6 | 2 (4.3) | | - | |
| 7 | 3 (6.4) | | - | |
| 8 | 5 (10.6) | | - | |
| Lost to follow-up | 1 (2.1) | | - | |
| **mRS (at 3 months)** | | | | |
| 0 | 1 (2.1) | | - | |
| 1 | 6 (12.8) | | - | |
| 2 | 8 (17.0) | | - | |
| 3 | 6 (12.8) | | - | |
| 4 | 5 (10.6) | | - | |
| 5 | 6 (12.8) | | - | |
| 6 | 14 (29.8) | | - | |
| Lost to follow-up | 1 (2.1) | | - | |

**Supplementary Table 3. Datasets**

Overview of the datasets used (provided along with the study).

| Dataset | Content | Filename |
|---|---|---|
| 1 | Proteomics (raw intensities) | dataset_1.csv |
| 2 | Proteomics (normalized intensities) | dataset_2.csv |
| 3 | Photospectrometric CSF measurements | dataset_3.csv |
| 4 | Clinical patient data | dataset_4.csv |
| 5 | TBARS from patient CSF (batch 1) | dataset_5.csv |
| 6 | TBARS from patient CSF (batch 2) | dataset_6.csv |
| 7 | TBARS Hb dose range | dataset_7.csv |
| 8 | Vascular function Hb dose range | dataset_8.csv |
| 9 | Vascular function Hb + Hp dose range | dataset_9.csv |
| 10 | Vascular function normalization plot | dataset_10.csv |
| 11 | Vascular function with patient CSF | dataset_11.csv |
